# The impact of parental adverse childhood experiences on children’s healthcare utilisation: a systematic review

**DOI:** 10.1101/2025.05.30.25328669

**Authors:** Mark F.M. Ketelaars, Merel Sprenger, Anouk J.M. Bos, Anne M. de Grauw, Michiel Burger, Mirjam van Veen, Jessica C. Kiefte-de Jong

**Affiliations:** Health Campus The Hague/Department of Public Health and Primary Care, Leiden University Medical Center, Turfmarkt 99, 2511 DP, The Hague, The Netherlands; Juliana Children’s Hospital (Haga Teaching hospital), Els Borst-Eilersplein 275, 2545 AA, The Hague, the Netherlands

**Keywords:** Adverse childhood experiences, intergenerational transmission, delivery of healthcare

## Abstract

**Background:** Adverse childhood experiences (ACEs) are linked to poor health and social outcomes, with growing interest in their intergenerational effects. While many studies have explored how ACEs affect adult health, the impact of parental ACEs on children’s healthcare utilisation has not yet been systematically reviewed.

**Methods:** We conducted a systematic review of original studies examining associations between parental ACEs or related adversity and children’s use of preventive, primary, or secondary healthcare. We searched PubMed, Embase, and PsycINFO up to December 2024. Title and abstract screening were conducted using ASReview. Quality of the studies was assessed using a composite framework evaluating study design, sampling, exposure and outcome measurement, and analysis strategy. Findings were synthesised narratively and visualised with harvest plots, grouped by healthcare level and exposure type.

**Results:** Out of 8494 records, 15 studies were included. Studies were heterogeneous in design, population, ACE measurement, and outcome definitions. No consistent associations were found in preventive or primary care domains. In secondary care, 11 of 12 associations were either positive or neutral; four high-quality studies showed increased use of emergency, inpatient, or psychiatric services among children of parents with ACEs. Methodological variability limited comparability between studies, particularly in how ACEs were operationalised.

**Conclusions:** Parental ACEs may contribute to increased use of secondary healthcare in offspring, though evidence across care levels remains inconclusive. Future studies should aim for more consistent measurement of ACEs and standardised outcome definitions to clarify intergenerational effects on healthcare utilisation.

**What is already known on this topic:**

- Adverse childhood experiences (ACEs) can have adverse effects on both mental and physical health. The last decade, more has become known about their intergenerational effects. For example, developmental and health effects have been observed in children of parents exposed to ACEs. Also, previous research has demonstrated the effects of ACEs on healthcare utilisation in adults. However, evidence for the effects of parental ACEs on healthcare utilisation of their children has not been systematically reviewed.

**What this study adds:**

- No association was found between parental ACEs and primary care utilisation, and mixed results were found for preventive care. However, a general trend suggests that parental ACEs increase secondary healthcare utilisation in children.

**How this study might affect research, practice or policy:**

- Higher secondary care utilisation in children of parents with ACEs implicates an increased burden of disease and added stress on healthcare systems. Addressing ACE-related healthcare challenges requires an integrated care approach that incorporates recognition and prevention of ACEs. In practice, this calls for early identification and coordinated family support. Future research should explore which care strategies best meet the needs of these families and how to effectively target upstream risk.

## Introduction

Adverse childhood experiences (ACEs) are potentially traumatic events that take place during childhood (0-17 years). Examples of these ACEs are child abuse or witnessing domestic violence, household substance abuse, neglect, psychological problems, and divorcing parents.^1^ A history of ACEs has been linked to a wide variety of problems, including chronic and mental illnesses, social cognitive impairments, and economic disadvantage.^2–5^ An accumulating body of literature provides growing evidence that ACEs can be transmitted across generations, with parents who have a history of ACEs potentially passing health effects on to their children.^5^ Several mechanisms are thought to play a role in this transmission, including impaired parenting resulting from the social-emotional effects of ACEs, epigenetic changes induced by early-life stress, and physiological changes associated with increased allostatic load, further increasing the risk of subsequent illnesses.^6,7^

The intergenerational transmission of ACEs is an important contributor to the overall burden of adversity on parents, children, healthcare, and society. Parental mental health conditions related to ACEs exacerbate long-term negative health outcomes and may increase healthcare use. Moreover, certain healthcare interventions themselves can become traumatic events.^8^ Additionally, Shah et al. describe that ACEs may lead to suboptimal coping abilities in parents that affect their capabilities to care for their child after hospitalisation.^9^ This interplay of adversity, caregiving strain, and systemic factors further increases the burden on healthcare and society, amplifying the direct effects of ACEs.

According to Hughes et al., ACEs are associated with major health issues and financial costs across European countries.^3^ However, the actual impact of parental ACEs on healthcare utilisation has never been systematically evaluated. Thus, more extensive research on the impact of parental ACEs and their healthcare consequences is needed for population health management purposes.^10^ Hence, this review aims to systematically examine the association between parental ACEs and children’s healthcare utilisation. Understanding this relationship can make way for reducing ACE-related complications in future generations and benefit both communities and patients.

## Methods

### DATA SOURCES AND SEARCH STRATEGY

This review followed a preregistered protocol (PROSPERO ID: 422942, accessible online via https://www.crd.york.ac.uk/PROSPERO/view/CRD42023422942) and PRISMA guidelines.^11^ We searched PubMed, Embase, and PsycINFO for literature on parental ACEs and healthcare utilisation published before December 2024, using terms related to ACEs, early life stress, and healthcare use (see online supplemental appendix 1). Only English and Dutch-language reports were included. Case reports, qualitative studies, and reviews were excluded. Grey literature was hand-searched via the DANS Data Station Life Sciences and British Library EThOS.

### ELIGIBILITY CRITERIA

Peer-reviewed studies assessing the association between parental ACEs and healthcare utilisation were eligible for inclusion. Studies had to meet two criteria: (1) the exposure involved parental, maternal, or paternal ACEs; and (2) the outcome described healthcare use or access by children. A broad definition of ACEs was used, allowing inclusion of studies using standardised tools (e.g. ACE Index, CTQ), individual events (e.g. physical violence, parental substance use), or less common adversities (e.g. migration). Healthcare utilisation included a wide range of services, such as GP visits, specialist care, well-child visits, dental care, treatment, and medication.

### STUDY SELECTION

All initially identified records were imported into Active Learning for Systematic Reviews (ASReview) for abstract screening. ASReview combines machine learning with active learning to predict inclusion likelihood.^12^ Based on prior evaluation studies, we selected the Naïve Bayes classifier with TF-IDF for feature extraction, and applied certainty-based sampling to prioritise relevant records. Screening was halted after 25% of records were reviewed, optimising efficiency while minimising irrelevant results.^13^

Subsequently, remaining records were imported into Mendeley for full-text screening using a standardised form (MS Forms) to assess eligibility and extract key information. Five reviewers (MK, JK, MS, AZ, AB) independently labelled studies as “included” or “excluded” with reasons; discrepancies were resolved through discussion. All inclusions were verified by the first author. Screening of reference lists from selected studies and prior reviews yielded one additional paper, resulting in 15 included studies. No separate list of excluded studies was compiled. Figure 1 shows the PRISMA flowchart.^11^

**Figure 1.**
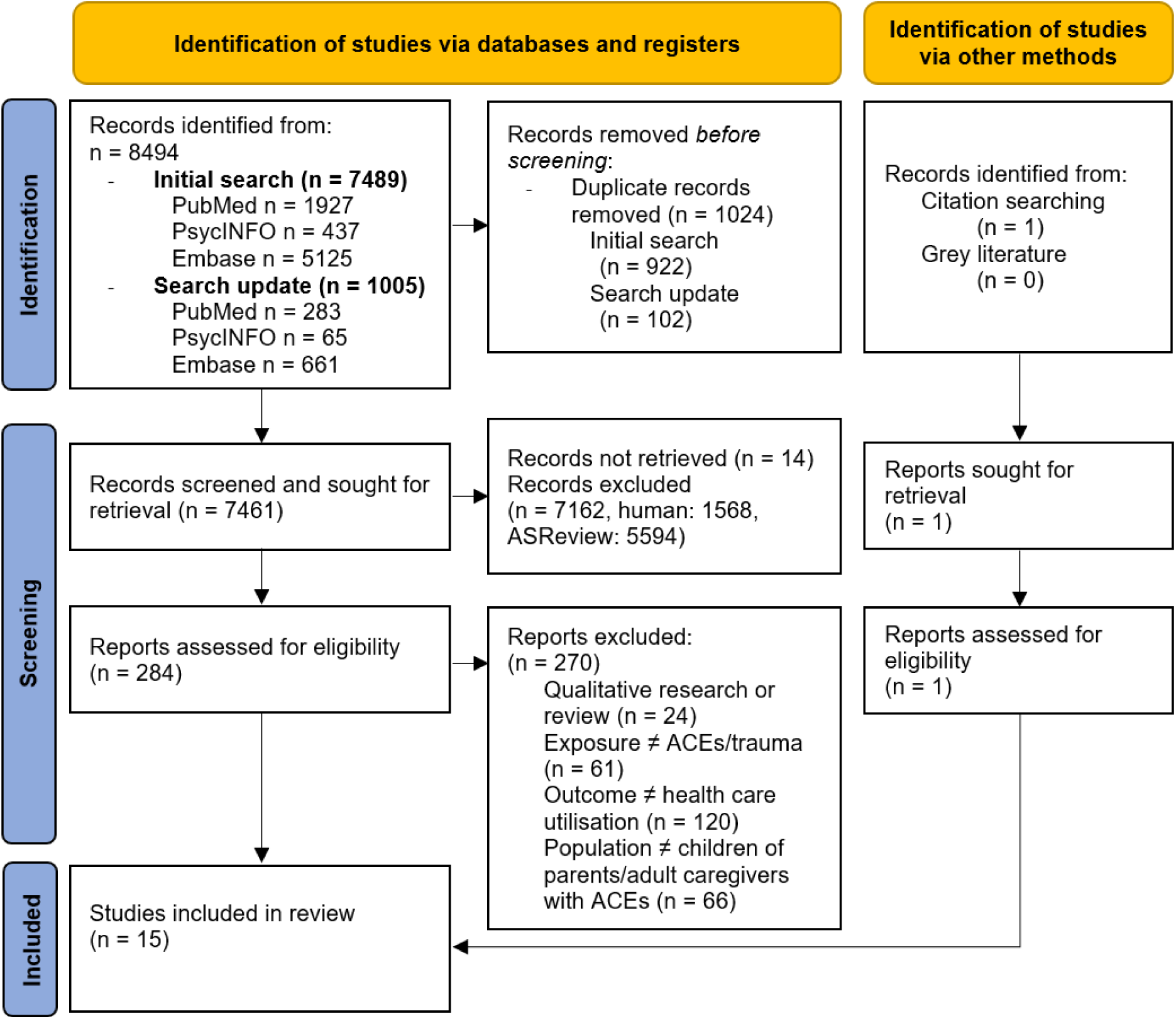
**PRISMA Flowchart.**

### DATA EXTRACTION

Data extraction was carried out by first author in accordance with the review protocol, on which a screening form and data extraction form were based (online supplemental appendix 2). Extracted data included article information, population characteristics (age, ethnicity and type of population), study design, ACE assessment, types of healthcare utilisation, and effect estimates.

### ASSESSMENT OF RISK OF BIAS

Study quality was assessed using a tool based on three established frameworks (online supplementary appendix 3).^14–16^ The checklist included five domains: study design, participant selection, outcome selection and measurement, and statistical analysis. Each domain was scored 0–2 points, except statistical analysis, which allowed up to 3 points for more rigorous confounder adjustment. Total scores ranged from 0 to 11 and were grouped into four categories: poor (0–3), fair (4–6), good (7–8), and excellent (9– 11). This approach provided a transparent and systematic method for comparing methodological quality across studies.

### DATA SYNTHESIS AND ANALYSIS

Results were synthesised in accordance with the SWiM reporting guidelines.^17^ For the 15 included papers, results were reported using harvest plots to visualise effects by study. Studies were ranked by quality score, and exposures and outcomes were categorised to summarise the heterogeneous findings. These groups were created after the inclusion of the studies, based on data availability. We created two groups for the exposures based on the ACE assessment: studies using the ACE index (ACE-10) as defined by Felitti et al., or studies using other assessments of parental adversity.^18^ Outcomes were grouped by levels of healthcare: preventive, primary and, secondary care. Since the healthcare levels are related in a hierarchical and integrated manner, this approach enables assessments of the mutual influence of utilisation in each of the levels. Within these groups, we further examined heterogeneity and certainty of findings by interpreting variation in ACE operationalisation, outcome definitions, population characteristics, and quality scores. The effect sizes reported in the studies were extracted without transformation. We did not convert the effect estimates to a standardised metric due to the heterogeneity in the studies. The results were synthesised by reporting the direction of effects, supported by metrics and confidence interval, where available. While all studies were included in the narrative synthesis, greater emphasis was placed on studies with higher methodological quality, validated ACE measures and well-defined healthcare utilisation outcomes.

## Results

Table 1 show the characteristics of the included studies. They were conducted between 2008 and 2024. The majority, 14 out of 15, were conducted in the United States. The total sample size across all studies was 106,775, with a median of 448 (range: 61–93,391). While only a small portion of studies reported parental age at time of the study, there was a wide range (15–50+ years), and mean maternal age was 34.4 years. The included publications studied children across all age groups (0–17 years), with a mean child age of 6.8 years. In most studies, the predominant population identified as Black. Most studies were cross-sectional observational. The mean study quality was 5.5 (SD 1.9), with a range of 3 to 9. The majority were rated as fair (61.9%), with only one (6.7%) rated as excellent. The findings are summarised in Figure 2, ordered by exposure and outcome domain and ranked by quality score. Studied exposures and outcomes were highly heterogeneous. The exposures, outcomes, and effect sizes are presented in Table 2.

**Figure 2.**
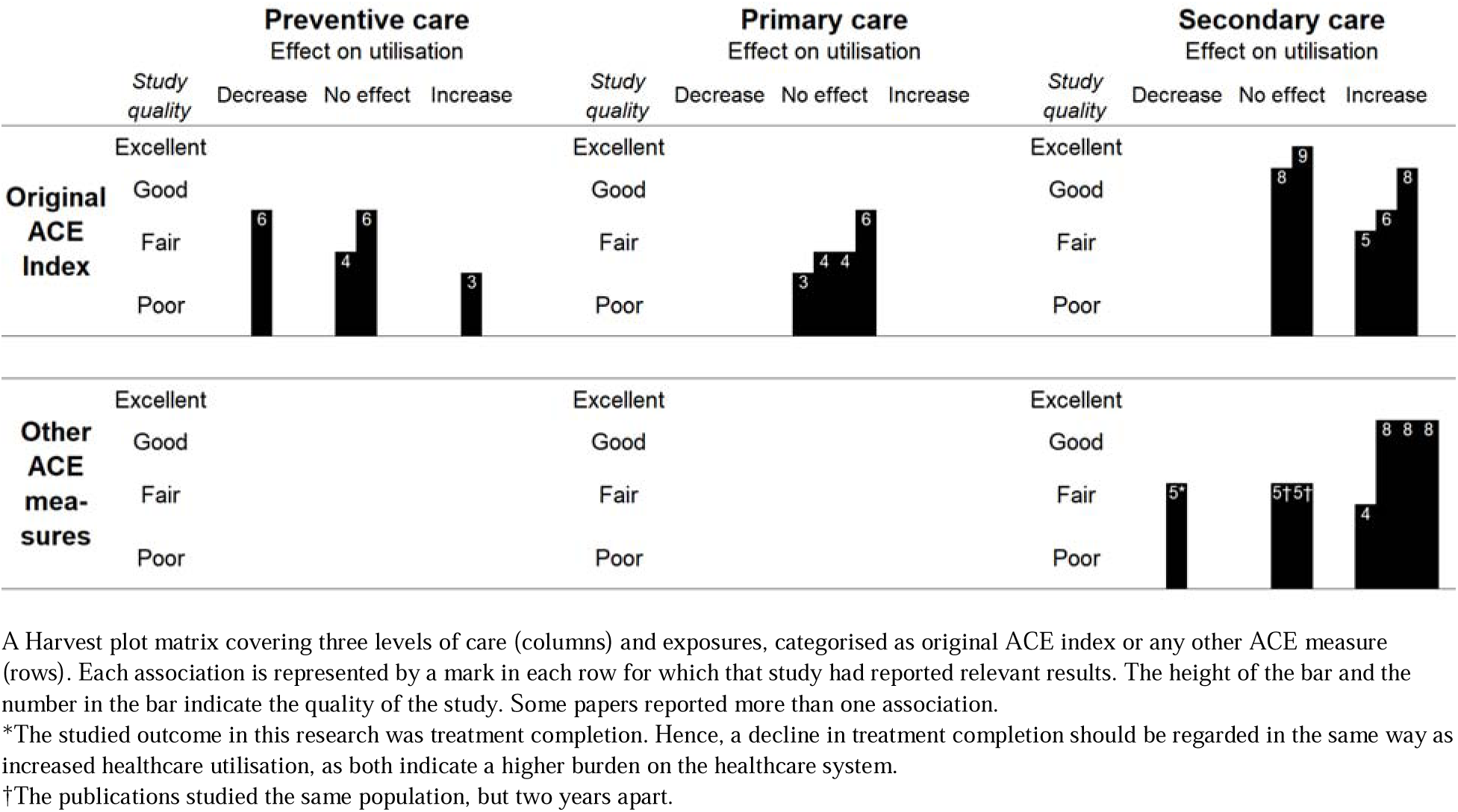
**Evidence for effects of parental ACEs on healthcare utilisation.**

**Table 1.**
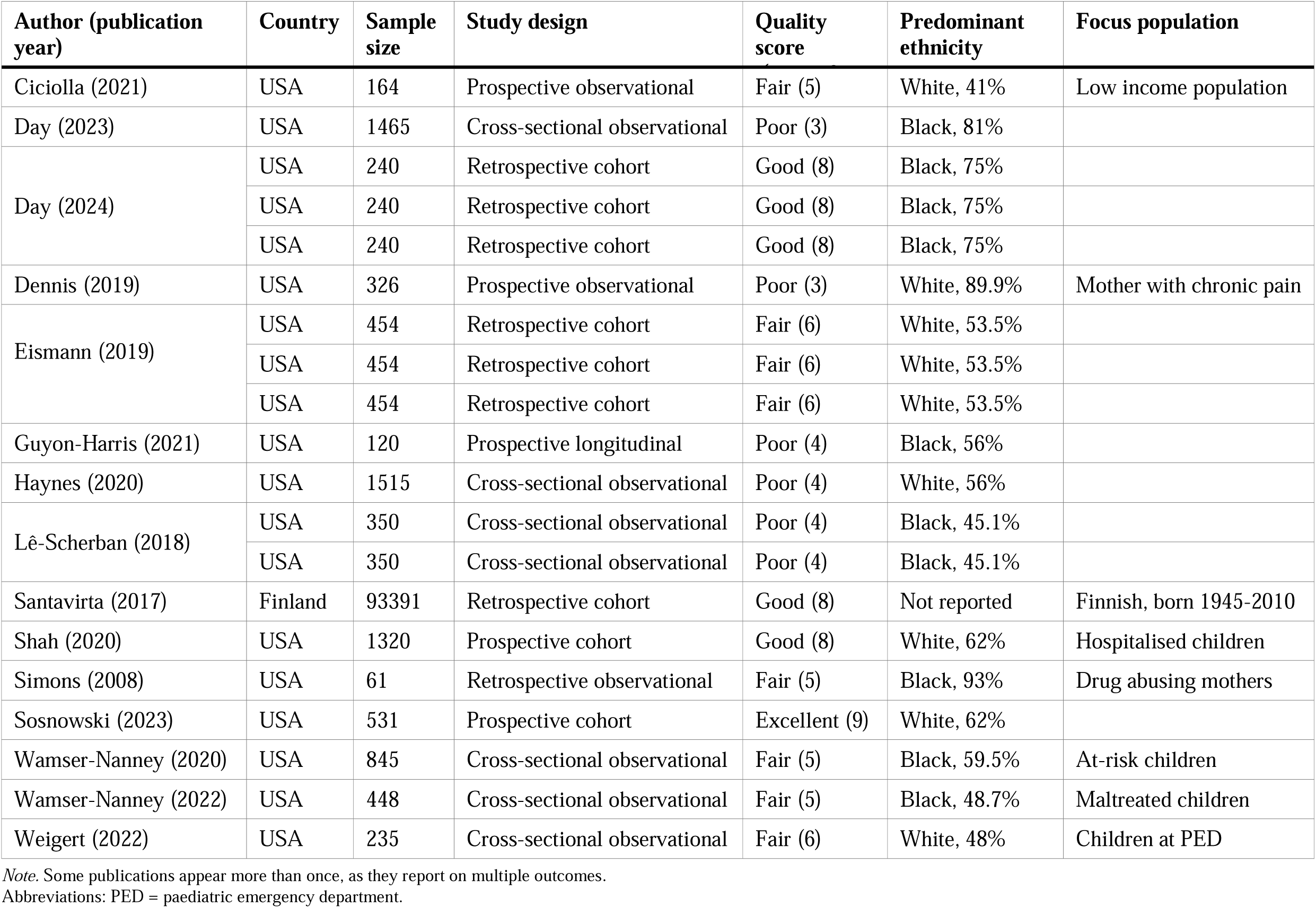
Overview of included study characteristics.

**Table 2.**
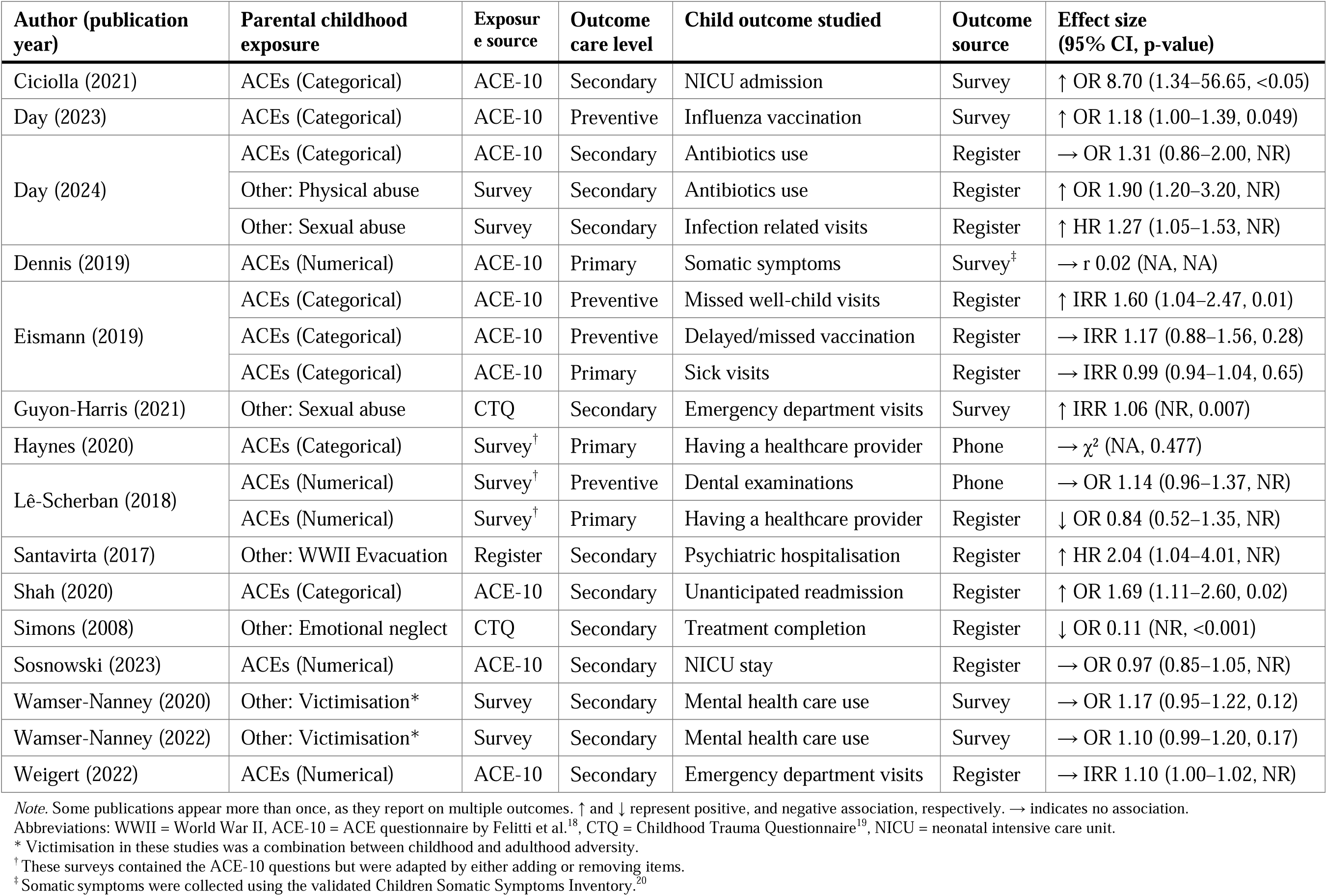
Overview all exposures, outcomes and effect sizes.

### PREVENTIVE CARE

Four preventive care outcomes were studied in three distinct reports, as Eismann et al. studied two outcomes. The outcomes assessed included vaccination uptake and well-child visit attendance. Mixed evidence was found in these studies, with some variation based on the type of exposure measured. Day et al. reported increased odds of paediatric influenza vaccination as caregiver ACEs increased (OR 1.18; 95% CI, 1.00–1.39, P = 0.049; quality score 3).^21^ Eismann et al. found that maternal, but not paternal, ACEs were associated with an increased probability of missing well-child visits (IRR 1.60; 95% CI, 1.04–2.47; P = 0.010; quality score 6).^22^ In the same study, they did not find an effect of parental ACEs on delayed or missed immunisations (as recommended by the American Academy of Pediatrics) (IRR 1.17; 95% CI, 0.88–1.56; P = 0.28; quality score 6). Lê-Scherban et al. found no association with dental examinations in the past twelve months (OR 1.14; 95% CI, 0.96–1.37, P = NR; quality score 4).^5^

There were no studies included in which exposures other than the original ACE index were studied in combination with preventive care services. Overall, findings on preventive care utilisation were mixed. Among the studies using categorical ACE exposures, results were inconsistent – one found a positive association, one negative, and one no association. The only study using a numerical ACE score reported no association with general preventive care. Two of the four studies had a low to fair study quality, which limits the certainty of findings in this domain. There were also some differences in the studied population: Eismann et al. reported predominantly white children (53.5%), Day reported predominantly Black children (81%) and Lê-Scherban reported 45.1% Black parents.

### PRIMARY CARE

Across primary care outcomes, none of the four studies found an association with parental ACEs. There were no studies with other exposures. The outcomes included sick visits or having a usual source of healthcare. In mothers with chronic pain, Dennis et al. found no correlation between maternal ACE scores and child somatic symptoms (r = 0.02; P = not reported, quality score 3). Eismann et al., using data from an urban paediatric clinic population, reported no association between parental ACEs and sick visits (IRR 0.99, 95% CI 0.94–1.04, P = 0.65; quality score 6). Haynes et al. found no relationship between categorical ACE scores and whether the child had a regular healthcare provider (P = 0.477; quality score 4). Finally, Lê-Scherban et al. found no association between parental ACE scores and having a usual source of care (OR 0.84, 95% CI 0.52–1.35, P = not reported; quality score 4).

Results were consistent in this domain; even though differences in population, quality score and methodology were observed, none of the studies found an association. The consistent absence of associations suggests limited evidence for a relationship of parental ACEs with paediatric healthcare utilisation. However, with three studies of fair quality and one of poor quality, the mean quality of studies in this domain (4.25) was lowest of all domains, so conclusions should be drawn with caution.

### SECONDARY CARE

Ten studies reported on secondary care utilisation, investigating twelve associations. Seven of these associations were positive, indicating increased healthcare utilisation for children of parents with ACEs; four showed no effect, and one indicated a decrease. Studied outcomes included both hospital related visits and treatment (completion). In the ACE index group, Shah et al. reported higher odds of unanticipated hospital readmission in children of parents with 4 or more ACEs (OR 1.69; 95% CI, 1.11– 2.60; quality score 8), Weigert et al. found a small increase in paediatric emergency department visits (IRR 1.013; 95% CI, 1.001–1.024; P = not reported; quality score 6) and Ciciolla et al. reported a sharp increase in odds of NICU admission for children born to mother with 6 more ACEs (OR 8.70; 95% CI, 1.34–56.65, P <0.05; quality score 6).^23–25^ Day et al. and Sosnowski et al. reported no association between parental ACEs and children’s antibiotic use and NICU stay, respectively.^26,27^

In the six studies reporting on other ACE measures, four positive associations were found. Day et al. reported an increase in antibiotics use in children of caregivers that were physically abused in their childhood – despite observing no association when ACEs were measured cumulatively (OR 1.90; 95% CI, 1.20–3.20, P = not reported; quality score 8). Moreover, in the same study, they found that caregiver childhood sexual abuse was associated with earlier infection-related clinical encounter (e.g., primary care, UC, ED, admission) in children (HR 1.27; 95% CI, 1.05–1.53, P = not reported).^26^ Additionally, Santavirta et al. found an increase in psychiatric hospitalisation in children whose mother had been evacuated from Finland during World War II (HR 2.04; 95% CI, 1.04–4.01, P = not reported; quality score 8) and Guyon-Harris et al. reported increased rates of infant emergency department visits when mothers had been sexually abused in childhood (IRR 1.06; 95% CI = not reported, P = 0.007, quality score 4).^28,29^ Wamser-Nanny et al. reported twice on a study in maltreated or at-risk children. They found a trend but no association between parental victimisation history and children’s mental healthcare use in 6-year-old maltreated children (OR 1.10, 95% CI 0.99–1.20, P = 0.07; quality score 5) and in 8-year-old at-risk children (OR 1.17, 95% CI 0.95–1.22, P = 0.12; quality score 5). In this study, parent victimisation combines childhood and adult trauma, hence, these results can’t be attributed to childhood adversity alone.^30,31^ Simons et al. reported the only negative association: drug-abusing mothers who had experienced emotional neglect and mental health issues were less likely to complete treatment programs with their children (OR 0.11; 95% CI = not reported, P <0.001; quality score 5).^32^ However, lower treatment completion could increase children’s disease burden, further supporting the trend from three high-quality studies that showed parental ACEs lead to increased secondary healthcare utilisation.

Findings in this domain were mixed, but most associations were positive. There was considerable heterogeneity in study design and outcomes. In the other ACE measure group, all but one study investigated a predominantly Black population. Despite these differences, the overall evidence tentatively points towards a link between parental ACEs and increased secondary care use, though this should be interpreted with caution.

## Discussion

Whilst our findings do not provide consistent associations across all healthcare domains, they suggest a positive trend for specific forms of healthcare utilisation. Parental ACEs showed no associations with primary care and yielded mixed results with preventive care, but a more consistent trend towards increased secondary care utilisation emerged.

To our knowledge, no other reviews have been published examining the association between parental ACEs and children’s healthcare utilisation. However, the trend towards increased secondary care utilisation is in line with existing literature, as Bhattarai reported increased all-cause hospitalisation in adults exposed to ≥1 ACE.^33^ In our review, 85% of studies examining hospital presentation or admission in children similarly reported an increase in utilisation. Together, these findings may reflect rising morbidity in children of parents with ACEs, with health problems potentially going unrecognised until symptoms escalate – resulting in greater reliance on acute rather than planned care.

The observed trend in secondary care, especially hospitalisation, may result from higher disease risk in children of parents with ACEs, combined with lower engagement with early healthcare services. Parents with ACEs may be unaware of their own or children’s increased health risks, as the intergenerational effects – potentially mediated by mechanisms such as allostatic load – are often gradual, biologically embedded, and hard to detect.^34^ Socioeconomic disadvantage and adversity-related behaviours – such as avoidance of care and reduced coping capacity – can further reduce timely healthcare seeking.^35^.^36–38^ Because of these factors, parents may feel less able or are less organised to come to visits as scheduled, which could be supported by the decrease found in treatment completion and increase in missed well-child visits in this review.^22,32,39^

Another explanation is a possible mismatch between the levels of care. Improved coordination between preventive, primary, and specialist services could help prevent unnecessary escalation of care – particularly in ACE-exposed individuals navigating complex care systems. The psychosocial needs underlying the reduced preventive care utilisation could be addressed by intervening at family level and early recognition of parental ACEs could help reduce disengagement or delayed care. This focus allows for targeting root causes of healthcare utilisation and improving long-term outcomes, while also prompting reflection on whether conventional service use metrics fully capture the support needs of these families. Barriers such as limited system trust, financial constraints, or use of alternative care may contribute to underutilisation of formal services, despite elevated health risks in children of parents with ACEs.

The findings in this research should be interpreted in light of some methodological considerations. First, although the quality of the included studies was generally fair to good, the applied analytics in these studies was highly heterogeneous, which precluded meta-analysis. Moreover, the SWiM-based synthesis revealed substantial methodological heterogeneity across studies, particularly in the operationalisation of parental ACEs. Some studies used numerical ACE scores, others categorical groupings, and multiple classification thresholds were applied. These differences complicate cross-study comparison and may influence effect estimates, with categorical approaches often yielding stronger associations. As noted in the results, where studies using categorical ACEs often produced stronger associations than numerical ones, this highlights the need for greater consensus on ACE operationalisation in future research.

Additional limitations include the possibility of publication and language bias, as only peer-reviewed English-language studies were included. Moreover, generalisability is limited: 14 of 15 studies were conducted in the United States, and many included ethnically specific or high-risk populations. This may limit the applicability of findings to broader or European contexts. Further research in diverse settings is needed to strengthen the evidence base and inform cross-context health policy.

While there is ample research on the association between ACEs and disease outcomes, evidence on how parental ACEs affect healthcare utilisation in children remains limited. Nonetheless, a trend towards increased secondary care utilisation was observed. This review highlights the need for further research to clarify these secondary consequences and to inform policy development. Investing in early identification and integrated, proactive care approaches may help reduce the intergenerational burden of ACEs on health systems and society.

## Ethics statements

### PATIENT CONSENT FOR PUBLICATION

Not applicable.

### ETHICS APPROVAL

Not applicable.

### Conflicts of interest

The authors declare no competing interests.

### Funding

No funding was received for the conduct of this review.

## Data Availability

All data produced in the present study are available upon reasonable request to the authors

## Acknowledgements

We thank Anne Zijp for her contribution to the screening process.

## Notes

### Competing Interest Statement

The authors have declared no competing interest.

### Funding Statement

This study did not receive any funding

### Summary of Updates

Updates based on editors remarks: more depth in the analysis and result section. Shortened the discussion section.

