## Supplementary material for "The impact of parental adverse childhood experiences on children’s healthcare utilisation: a systematic review": online supplemental appendix

Supplementary appendix 1. Search terms.

PUBMED

((("Parents"[MeSH] OR "parent*"[tw] OR "father*"[tw] OR "mother*"[tw] OR "stepfather*"[tw] OR "stepmother*"[tw]) AND ("Adverse Childhood Experiences"[MeSH] OR "Adverse Childhood Experience*"[tw] OR "Adverse Childhood Event*"[tw] OR "Childhood Trauma*"[tw] OR "Traumatic Childhood Experience*"[tw] OR "Adolescent Trauma*"[tw] OR "Early Life Stress*"[tw] OR "Early-Life Stress*"[tw] OR (Adverse[ti] AND Childhood*[ti] AND Experience*[ti]) OR (childhood*[ti] AND trauma*[ti]) OR (adolescen*[ti] AND trauma*[ti]) OR (("early life"[ti] OR "early-life"[ti]) AND stress*[ti]) OR ("Adverse Childhood"[title/abstract:~2] AND ("Childhood experience"[title/abstract:~2] OR "Childhood experiences"[title/abstract:~2] OR "adverse experience"[title/abstract:~2] OR "adverse experiences"[title/abstract:~2])) OR ("Adverse Childhood"[title/abstract:~2] AND ("Childhood event"[title/abstract:~2] OR "Childhood events"[title/abstract:~2] OR "adverse event"[title/abstract:~2] OR "adverse events"[title/abstract:~2])) OR "Childhood Trauma"[title/abstract:~1] OR "Childhood Traumas"[title/abstract:~1] OR "Childhood Traumatic"[title/abstract:~1] OR "adolescent trauma"[title/abstract:~1] OR "adolescent traumas"[title/abstract:~1] OR "adolescent traumatic"[title/abstract:~1] OR "adolescents trauma"[title/abstract:~1] OR " adolescents traumas"[title/abstract:~1] OR " adolescents traumatic"[title/abstract:~1] OR "adolescence trauma"[title/abstract:~1] OR "adolescence traumas"[title/abstract:~1] OR "adolescence traumatic"[title/abstract:~1] OR "early life stress"[title/abstract:~1] OR "early-life stress"[title/abstract:~1] OR "early life stressfull"[title/abstract:~1] OR "early-life stressful"[title/abstract:~1] ) AND ("Delivery of Health Care"[MeSH] OR "Health Care Quality, Access, and Evaluation"[Mesh:NoExp] OR "Health Services Research"[Mesh] OR "Health Services"[mesh] OR "Health service*"[tw] OR "Public Health Systems Research"[Mesh] OR "Quality of Health Care"[Mesh] OR "Delivery of Health Care"[tw] OR "Delivery of HealthCare"[tw] OR "Delivery of Care"[tw] OR "Quality of Health Care"[tw] OR "Quality of Healthcare"[tw] OR "Quality of care"[tw] OR "health care quality"[tw] OR "healthcare quality"[tw] OR "care quality"[tw] OR "Health Care Utilis*"[tw] OR "Health Care Utiliz*"[tw] OR "Health Care seek*"[tw] OR "Health Care Us*"[tw] OR "Healthcare Utilis*"[tw] OR "Healthcare Utiliz*"[tw] OR "Healthcare seek*"[tw] OR "seeking health care"[tw] OR "seeking healthcare"[tw] OR "seek health care"[tw] OR "seek healthcare"[tw] OR "seeked health care"[tw] OR "seeked healthcare"[tw] OR "Health care Us*"[tw] OR "Use health care"[tw] OR "Usage health care"[tw] OR "using health care"[tw] OR "Healthcare Us*"[tw] OR "Use of healthcare"[tw] OR "Usage of healthcare"[tw] OR "using health care"[tw] OR "Health Care Utilisation"[title/abstract:~1] OR "Health Care Utilisations"[title/abstract:~1] OR "Health Care Utilised"[title/abstract:~1] OR "Health Care Utilization"[title/abstract:~1] OR "Health Care Utilizations"[title/abstract:~1] OR "Health Care Utilized"[title/abstract:~1] OR "Health Care seek"[title/abstract:~1] OR "Health Care seeking"[title/abstract:~1] OR "Health Care seeks"[title/abstract:~1] OR "Health Care seeked"[title/abstract:~1] OR "Health Care Use"[title/abstract:~1] OR "Health Care Uses"[title/abstract:~1] OR "Health Care Used"[title/abstract:~1] OR "Health Care Using"[title/abstract:~1] OR "Healthcare Utilisation"[title/abstract:~1] OR "Healthcare Utilisations"[title/abstract:~1] OR "Healthcare Utilised"[title/abstract:~1] OR "Healthcare Utilization"[title/abstract:~1] OR "Healthcare Utilizations"[title/abstract:~1] OR "Healthcare Utilized"[title/abstract:~1] OR "Healthcare seek"[title/abstract:~1] OR "Healthcare seeking"[title/abstract:~1] OR "Healthcare seeks"[title/abstract:~1] OR "Healthcare seeked"[title/abstract:~1] OR "Healthcare Use"[title/abstract:~1] OR "Healthcare Uses"[title/abstract:~1] OR "Healthcare Used"[title/abstract:~1] OR "Healthcare Using"[title/abstract:~1] OR " Care Utilis*"[tw] OR " Care Utiliz*"[tw] OR " Care seek*"[tw] OR " Care Us*"[tw] OR "care Utilis*"[tw] OR "care Utiliz*"[tw] OR "care seek*"[tw] OR "seeking care"[tw] OR "seeking care"[tw] OR "seek care"[tw] OR "seek care"[tw] OR "seeked care"[tw] OR "seeked care"[tw] OR "care Us*"[tw] OR "Use care"[tw] OR "Usage care"[tw] OR "using care"[tw] OR "care Us*"[tw] OR "Use of care"[tw] OR "Usage of care"[tw] OR "using care"[tw] OR " Care Utilisation"[title/abstract:~1] OR " Care Utilisations"[title/abstract:~1] OR " Care Utilised"[title/abstract:~1] OR " Care Utilization"[title/abstract:~1] OR " Care Utilizations"[title/abstract:~1] OR " Care Utilized"[title/abstract:~1] OR " Care seek"[title/abstract:~1] OR " Care seeking"[title/abstract:~1] OR " Care seeks"[title/abstract:~1] OR " Care seeked"[title/abstract:~1] OR " Care Use"[title/abstract:~1] OR " Care Uses"[title/abstract:~1] OR " Care Used"[title/abstract:~1] OR " Care Using"[title/abstract:~1] OR "care Utilisation"[title/abstract:~1] OR "care Utilisations"[title/abstract:~1] OR "care Utilised"[title/abstract:~1] OR "care Utilization"[title/abstract:~1] OR "care Utilizations"[title/abstract:~1] OR "care Utilized"[title/abstract:~1] OR "care seek"[title/abstract:~1] OR "care seeking"[title/abstract:~1] OR "care seeks"[title/abstract:~1] OR "care seeked"[title/abstract:~1] OR "care Use"[title/abstract:~1] OR "care Uses"[title/abstract:~1] OR "care Used"[title/abstract:~1] OR "care Using"[title/abstract:~1] OR "Somatoform Disorders"[Mesh] OR "Somatoform Disorders"[title/abstract:~1] OR "Somatoform Disorder"[title/abstract:~1] OR "somatic disorder"[title/abstract:~1] OR "somatic disorders"[title/abstract:~1] OR "functional disorder" [title/abstract:~1] OR "functional disorders" [title/abstract:~1] OR (seek*[ti] AND help[ti]) OR "seek help"[title/abstract:~1] OR "seeks help"[title/abstract:~1] OR "seeked help"[title/abstract:~1] OR "seeking help"[title/abstract:~1] OR ("later life"[title/abstract:~4] AND ("Therapeutics"[mesh] OR "therapy"[tw] OR "therapies"[tw] OR " therapeutics"[tw] OR "therapeutic"[tw] OR "treatment"[tw] OR "treatments"[tw] OR "treated"[tw] OR "treat"[tw] OR "treats"[tw] OR "treating"[tw])) OR "Costs and Cost Analysis"[Mesh] OR cost[tw] OR costs[tw] OR burden[tw] OR burdens[tw] OR "Primary Health Care"[Mesh] OR "primary health care"[tw] OR "primary healthcare"[tw] OR "primary care"[tw] OR "secondary health care"[tw] OR "secondary healthcare"[tw] OR "secondary care"[tw] OR "Hospitalization"[Mesh] OR hospitalizat*[tw] OR hospitalisat*[tw] OR admission*[tw] OR ("number visits"[title/abstract:~2] OR "numbers visits"[title/abstract:~2]))) NOT Review[Publication Type] NOT "Systematic Review"[Publication Type] NOT "Case Reports"[Publication Type] NOT Editorial[Publication Type] NOT Letter[Publication Type] NOT "Animals"[Tiab]) AND (DUTCH[Language] OR ENGLISH[Language]) NOT ("Animals"[Mesh] NOT ("Animals"[Mesh] AND "Humans"[Mesh]))

EMBASE

1	(exp parent/ or "parent*".mp. or "father*".mp. or "mother*".mp. or "stepfather*".mp. or "stepmother*".mp.) and (exp childhood adversity/ or "Adverse Childhood Experience*".mp. or "Adverse Childhood Event*".mp. or "Childhood Trauma*".mp. or "Traumatic Childhood Experience*".mp. or "Adolescent Trauma*".mp. or "Early Life Stress*".mp. or "Early-Life Stress*".mp. or (Adverse and Childhood* and Experience*).ti. or (childhood* and trauma*).ti. or (adolescen* and trauma*).ti. or (("early life" or "early-life") and stress*).ti. or ((Adverse adj2 Childhood) and ((Childhood adj2 experience*) or (adverse adj2 experience*))).mp. or ((Adverse adj2 Childhood) and ((Childhood adj2 event*) or (adverse adj2 event*))).mp. or (Childhood adj2 Trauma*).mp. or (adolescen* adj2 trauma*).mp. or (early life adj2 stress).mp. or (early life adj2 stressfull).mp.) and (exp health care delivery/ or health care quality/ or exp health services research/ or exp "health service"/ or "health service*".mp. or exp public health systems research/ or "Delivery of Health Care".mp. or "Delivery of HealthCare".mp. or "Delivery of Care".mp. or "Quality of Health Care".mp. or "Quality of Healthcare".mp. or "Quality of care".mp. or "health care quality".mp. or "healthcare quality".mp. or "care quality".mp. or "Health Care Utilis*".mp. or "Health Care Utiliz*".mp. or "Health Care seek*".mp. or "Health Care Us*".mp. or "Healthcare Utilis*".mp. or "Healthcare Utiliz*".mp. or "Healthcare seek*".mp. or "seeking health care".mp. or "seeking healthcare".mp. or "seek health care".mp. or "seek healthcare".mp. or "seeked health care".mp. or "seeked healthcare".mp. or "Health care Us*".mp. or "Use health care".mp. or "Usage health care".mp. or "using health care".mp. or "Healthcare Us*".mp. or "Use of healthcare".mp. or "Usage of healthcare".mp. or "using health care".mp. or (Health Care adj1 Utili*).mp. or (Health Care adj1 seek*).mp. or (Health Care adj1 Use*).mp. or (Health Care adj1 Using).mp. or (Healthcare adj1 Utili*).mp. or (Healthcare adj1 seek*).mp. or (Healthcare adj1 Utili*).mp. or (Healthcare adj1 seek*).mp. or (Healthcare adj1 Use*).mp. or (Healthcare adj1 Using).mp. or "Care Utilis*".mp. or "care Utiliz*".mp. or "care seek*".mp. or "care us*".mp. or "care utilis*".mp. or "care utiliz*".mp. or "care seek*".mp. or "seeking care".mp. or "seeking care".mp. or "seek care".mp. or "seek care".mp. or "seeked care".mp. or "seeked care".mp. or "care us*".mp. or "Use care".mp. or "usage care".mp. or "using care".mp. or "care us*".mp. or "use of care".mp. or "usage of care".mp. or "using care".mp. or (Care adj1 utili*).mp. or (Care adj1 seek*).mp. or (Care adj1 Use*).mp. or (Care adj1 Using).mp. or exp somatoform disorder/ or (Somatoform adj1 Disorder*).mp. or (somatic adj1 disorder*).mp. or (functional adj1 disorder*).mp. or (seek* and help).ti. or (seek* adj1 help).mp. or ((later adj4 life).mp. and (exp therapy/ or therapy.mp. or therapies.mp. or therapeutics.mp. or therapeutic.mp. or treatment.mp. or treatments.mp. or treated.mp. or treat.mp. or treats.mp. or treating.mp.)) or exp cost/ or cost.mp. or costs.mp. or burden.mp. or burdens.mp. or exp primary health care/ or "primary health care".mp. or "primary healthcare".mp. or "primary care".mp. or "secondary health care".mp. or "secondary healthcare".mp. or "secondary care".mp. or exp hospitalization/ or hospitalizat*.mp. or hospitalisat*.mp. or admission*.mp. or (number* adj2 visits).mp.) [mp=title, abstract, heading word, drug trade name, original title, device manufacturer, drug manufacturer, device trade name, keyword heading word, floating subheading word, candidate term word]	7046

2	limit 1 to ("systematic review" and ("reviews (maximizes sensitivity)" or "reviews (maximizes specificity)" or "reviews (best balance of sensitivity and specificity)") and (editorial or letter or "review"))	83

3	1 not 2	6963

4	limit 3 to (dutch or english)	6611

5	limit 4 to "humans only (removes records about animals)"	6516

6	limit 1 to "article"	5125

PSYCINFO

(DE "Parents" OR DE "Adoptive Parents" OR DE "Fathers" OR DE "Foster Parents" OR DE "Homosexual Parents" OR DE "Mothers" OR DE "Single Parents" OR DE "Stepparents" OR TX "parent*" OR TX "father*" OR TX "mother*" OR TX "stepfather*" OR TX "stepmother*") AND (DE "Childhood Adversity" OR TX "Adverse Childhood Experience*" OR TX "Adverse Childhood Event*" OR TX "Childhood Trauma*" OR TX "Traumatic Childhood Experience*" OR TX "Adolescent Trauma*" OR TX "Early Life Stress*" OR TX "Early-Life Stress*" OR (TI Adverse AND TI Childhood* AND TI Experience*) OR (TI childhood* AND TI trauma*) OR (TI adolescen* AND TI trauma*) OR ((TI "early life") AND TI stress*) OR ((Adverse NEAR3 Childhood) AND ((Childhood NEAR3 experience*) OR (adverse NEAR3 experience*))) OR ((Adverse NEAR3 Childhood) AND ((Childhood NEAR3 event*) OR (adverse NEAR3 event*))) OR (Childhood NEAR3 Trauma*) OR (adolescen* NEAR3 trauma*) OR (early life NEAR3 stress) OR (early life NEAR3 stressfull)) AND (DE "Health Care Delivery" OR DE "Clinical Practice" OR DE "Health Care Access" OR DE "mental health services" OR DE "Health Care Costs" OR DE "Health Care Reform" OR DE "Health Care Utilization" OR DE "Managed Care" OR DE "Quality of Care" OR DE "Quality of Services" OR DE "Public Health Research" OR TX "Delivery of Health Care" OR TX "Delivery of HealthCare" OR TX "Delivery of Care" OR TX "Quality of Health Care" OR TX "Quality of Healthcare" OR TX "Quality of care" OR TX "health care quality" OR TX "healthcare quality" OR TX "care quality" OR TX "health service*" OR TX "Health Care Utilis*" OR TX "Health Care Utiliz*" OR TX "Health Care seek*" OR TX "Health Care Us*" OR TX "Healthcare Utilis*" OR TX "Healthcare Utiliz*" OR TX "Healthcare seek*" OR TX "seeking health care" OR TX "seeking healthcare" OR TX "seek health care" OR TX "seek healthcare" OR TX "seeked health care" OR TX "seeked healthcare" OR TX "Health care Us*" OR TX "Use health care" OR TX "Usage health care" OR TX "using health care" OR TX "Healthcare Us*" OR TX "Use of healthcare" OR TX "Usage of healthcare" OR TX "using health care" OR (Health Care NEAR2 Utili*) OR (Health Care NEAR2 seek*) OR (Health Care NEAR2 Use*) OR (Health Care NEAR2 Using) OR (Healthcare NEAR2 Utili*) OR (Healthcare NEAR2 seek*) OR (Healthcare NEAR2 Utili*) OR (Healthcare NEAR2 seek*) OR (Healthcare NEAR2 Use*) OR (Healthcare NEAR2 Using) OR TX "Care Utilis*" OR TX "care Utiliz*" OR TX "care seek*" OR TX "care us*" OR TX "care utilis*" OR TX "care utiliz*" OR TX "care seek*" OR TX "seeking  care" OR TX "seeking care" OR TX "seek care" OR TX "seek care" OR TX "seeked care" OR TX "seeked care" OR TX "care us*" OR TX "Use  care" OR TX "usage  care" OR TX "using  care" OR TX "care us*" OR TX "use of care" OR TX "usage of care" OR TX "using  care" OR (Care NEAR2 utili*) OR (Care NEAR2 seek*) OR (Care NEAR2 Use*) OR (Care NEAR2 Using) OR DE "Somatoform Disorders" OR DE "Body Dysmorphic Disorder" OR DE "Conversion Disorder" OR DE "Factitious Disorders" OR DE "Illness Anxiety Disorder" OR DE "Neurasthenia" OR DE "Somatization Disorder" OR DE "Somatoform Pain Disorder" OR (Somatoform NEAR2 Disorder*) OR (somatic NEAR2 disorder*) OR (functional NEAR2 disorder*) OR (TI seek* AND TI help) OR (seek* NEAR2 help) OR ((later NEAR5 life) AND (DE "Treatment" OR DE "Addiction Treatment" OR DE "Adjunctive Treatment" OR DE "Adventure Therapy" OR DE "Aftercare" OR DE "Alternative Medicine" OR DE "Anxiety Management" OR DE "Behavior Modification" OR DE "Bibliotherapy" OR DE "Brief Interventions" OR DE "Caregiving" OR DE "Client Transfer" OR DE "Client Treatment Matching" OR DE "Cognitive Behavior Therapy" OR DE "Cognitive Stimulation Therapy" OR DE "Cognitive Techniques" OR DE "Computer Assisted Therapy" OR DE "Counseling" OR DE "Creative Arts Therapy" OR DE "Cross Cultural Treatment" OR DE "Culturally Adapted Interventions" OR DE "Disease Management" OR DE "Habilitation" OR DE "Health Care Services" OR DE "Horticulture Therapy" OR DE "Hospice" OR DE "Human Potential Movement" OR DE "Human Services" OR DE "Hydrotherapy" OR DE "Institutionalization" OR DE "Integrated Services" OR DE "Interdisciplinary Treatment Approach" OR DE "Intervention" OR DE "Involuntary Treatment" OR DE "Language Therapy" OR DE "Life Sustaining Treatment" OR DE "Maintenance Therapy" OR DE "Medical Treatment (General)" OR DE "Mental Health Programs" OR DE "Milieu Therapy" OR DE "Mind Body Therapy" OR DE "Mindfulness-Based Interventions" OR DE "Movement Therapy" OR DE "Multimodal Treatment Approach" OR DE "Multisystemic Therapy" OR DE "Outpatient Treatment" OR DE "Pain Management" OR DE "Partial Hospitalization" OR DE "Personal Therapy" OR DE "Physical Treatment Methods" OR DE "Private Practice" OR DE "Psychoeducation" OR DE "Psychotherapy" OR DE "Rehabilitation" OR DE "Relaxation Therapy" OR DE "Respite Care" OR DE "Self-Help Techniques" OR DE "Sex Therapy" OR DE "Social Casework" OR DE "Sociotherapy" OR DE "Speech Therapy" OR DE "Spiritual Care" OR DE "Strengths-Based Interventions" OR DE "Symptoms Based Treatment" OR DE "Therapeutic Processes" OR DE "Transdiagnostic Treatment" OR DE "Trauma-Informed Care" OR DE "Trauma Treatment" OR DE "Treatment Guidelines" OR DE "Treatment Outcomes" OR DE "Treatment Planning" OR DE "Video-Based Interventions" OR TX therapy OR TX therapies OR TX therapeutics OR TX therapeutic OR TX treatment OR TX treatments OR TX treated OR TX treat OR TX treats OR TX treating)) OR DE "Costs and Cost Analysis" OR DE "Budgets" OR DE "Cost Containment" OR DE "Health Care Costs" OR DE "Money" OR DE "Health Care Reimbursement" OR DE "Insurance" OR DE "Health Insurance" OR DE "Life Insurance" OR DE "Social Security" OR DE "Professional Fees" OR DE "Fee for Service" OR TX cost OR TX costs OR TX burden OR TX burdens OR DE "Primary Health Care" OR TX "primary health care" OR TX "primary healthcare" OR TX "primary care" OR TX "secondary health care" OR TX "secondary healthcare" OR TX "secondary care" OR DE "Hospitalization" OR DE "Hospital Admission" OR DE "Hospital Discharge" OR DE "Mental Health Commitment" OR DE "Psychiatric Hospitalization" OR TX hospitalizat* OR TX hospitalisat* OR TX admission* OR (number* NEAR3 visits))

**Limiters Applied**
Language: Dutch, English; Population Group: Human, Male, Transgender, Female, Inpatient, Outpatient; Document Type: Abstract Collection, Bibliography, Chapter, Clarification, Column/Opinion, Comment/Reply, Dissertation, Encyclopedia Entry, Erratum/Correction, Interview, Journal Article, Obituary, Poetry, Publication Information, Reprint, Retraction

XLanguage: Dutch, English

XPopulation Group: Human

XDocument Type: Journal Article

Supplementary appendix 2. Screening form. Adapted from Microsoft Forms.

Hello! Thanks for helping me out with the selection of papers for my review.

**Some instructions**: please follow the link to Mendeley to find your own folder in the shared group. This folder contains all papers that were selected for you. Double click the reference and Mendeley should open the pdf for you.

Please make sure you are working in the right folder. You should be able to find your own folder as in this example. Start in the "All assigned" folder and copy a paper to either excluded or included.

1. Author of the paper you are screening (last name of first author)
2. Publication year of the paper you are screening
3. Full title of the paper you are screening

**Exclusion criteria**

If any of the following is applicable to the paper, it should be excluded:

- The body of evidence is written in any other language than English or Dutch
- Case reports and reviews
- Qualitative research

☐ The body of evidence is written in any other language than English or Dutch
☐ Paper is a case report or review
☐ Paper describes qualitative research
☐ None of the above

**Exposure: ACEs**

There are numerous types of potential traumatic events during childhood. In the literature, researchers work with many types and indexes. In 1998, Felitti et al. introduced the term Adverse Childhood Experiences and included 10 types of abuse in their index. We refer to this index as the original ACE index.

**The original ACE index consists of:**

- Emotional Abuse
- Physical Abuse
- Sexual Abuse
- Emotional Neglect
- Physical Neglect
- Parental Separation or Divorce
- Mother treated violently
- Household Substance Abuse
- Household Mental Illness
- Incarcerated Household Member

**Additional ACE items include:**

- Spanking
- Parental Gambling
- Foster care or child placing organisations
- Poverty
- Peer victimisation
- Neighbourhood safety

**Does the paper describe any type of ACE or traumatic event during childhood as the exposure?**
☐ Yes
☐ No

**Which of the following best describes the exposure?**
☐ Adverse Childhood Experiences (original index)
☐ [Tick individual ACEs and/or list others under “Andere”]

**Outcome: healthcare usage**

Healthcare usage can comprise the following sub-outcomes:

- Health care usage in general
- Primary and secondary care
- Treatment
- Health care costs
- Hospital admissions
- Functional disorders
- Persistent somatic disorders

(Note: diagnoses are not part of health care utilisation.)

☐ No healthcare utilisation outcome
☐ Health care usage in general
☐ Primary care including general practitioners
☐ Secondary care including medical specialists
☐ Hospital admissions
☐ Treatment
☐ Health care costs
☐ Functional disorders
☐ Persistent somatic disorders
☐ Dental care
☐ Foster care
☐ Andere

**Population**

This review is focussed on healthcare utilisation related to the intergenerational transmission of ACEs. Hence, the desired population are parents and their children. However, given the lack of research on ACEs and healthcare utilisation, looking only at parents and children might be too specific. Therefore, we will also include studies with adults alone (regardless of being a parent) and children alone (regardless of their parents).

If a specific patient population (e.g., children with asthma or diabetes patients) was targeted, please describe this population in the “other” field.

☐ Adults
☐ Children
☐ Children of parents/adult caregivers with ACEs
☐ Parents/adult caregivers with ACEs
☐ Andere

**Include or exclude?**

After answering the following question(s), please flag paper from your own folder 'To do' folder to either the 'Included' or 'Excluded' folder. You can then delete the paper from the 'To do' folder in order to get a clear overview of your progress.

☐ Yes, include this paper
☐ No, exclude this paper

**Please give the main reason(s) for exclusion**

Supplementary appendix 3. Quality score for selected studies.

Quality score
Systematic review: Parental ACEs – Children’s health care utilisation

This quality score was used to assess the quality of included studies in this systematic review and is applicable to both interventional and observational studies. The score was designed based on previously published scoring systems (1, 2). The quality score consists of 5 items, and each item is allocated 0, 1 or 2 points. This allows a total score between 0 and 10 points, 10 representing the highest quality.

1. Study design

**0** for studies with cross-sectional data collection

**1** for studies with longitudinal data collection or cohort studies

2. Study size

Observational studies

**0** if n<500

**1** if n 500 to 2000

**2** if >2000

3. Exposure

Observational studies

**0** if the study used no appropriate assessment or if not reported

**1** if the study used an appropriate assessment but not validated for the particular population

**1** if the study used an appropriate assessment but representing a single type of abuse rather than ACEs

**2** if the study used a validated assessment.

4. Outcome

**0** if the study used no appropriate outcome measurement method or if not reported

**1** if the study used a self-reported outcome measurements

**2** if the study used an objective and validated measurements

5. Adjustments

**0** if findings are not controlled for at least the two key covariates† mentioned below

**1** if findings are controlled for the following key confounders: age, sex, socioeconomic status

**2** if adequately randomized or if findings are additionally controlled for at least two of the following covariates†: age, sex, socioeconomic status, parent mental health, employment status, parental health conditions, child health conditions

**3** If effect modifications or stratifications for previous diagnoses, developmental problems or behavioural problems were taken into account in the analysis

† Either adjusted for in the statistical analyses; stratified for in the analyses; or not applicable (e.g. a study within a specific population does not require controlling for country of birth).

Note: this quality score is based on three existing scoring frameworks:

- Stroup DF, Berlin JA, Morton SC, Olkin I, Williamson GD, Rennie D, Moher D, Becker BJ, Sipe TA, Thacker SB. Meta-analysis of obser national studies in epidemiology: a proposal for reporting. Meta analysis Of Observational Studies in Epidemiology (MOOSE) group. JAMA 2000;283:2008–12.
- von Elm E, Altman DG, Egger M, Pocock SJ, Gotzsche PC, Vandenbroucke JP. Strobe Initiative. The Strengthening the Reporting of Observational Studies in Epidemiology (STROBE) statement: guidelines for reporting observational studies. J Clin Epidemiol 2008; 61:344–9.
- Wong WC, Cheung CS, Hart GJ. Development of a quality assess ment tool for systematic reviews of observational studies (QATSO) of HIV prevalence in men having sex with men and associated risk behaviours. Emerg Themes Epidemiol 2008;5:23.
